# Incidence of cardiovascular-kidney-metabolic syndrome and its risk factors for progression in China

**DOI:** 10.1101/2024.08.07.24311650

**Authors:** Aomiao Chen, Qiuyu He, Yichuan Wu, Jiaqi Chen, Xiaoqin Ma, Geningyue Wang, Lingyuan Hu, Zhuotong Wang, Jinming Huang, Xinran Xie, Yaoming Xue, Zongji Zheng, Yijie Jia

## Abstract

**Background:** Cardiovascular-kidney-metabolic syndrome (CKM syndrome) has become one of the leading causes of death. However, its prevalence and factors associated with its progression are unknown. In this study, we investigate the incidence of CKM syndrome in middle-aged and elderly individuals, identify the risk factors for CKM syndrome progression via 4 years of follow-up data, explore CKM syndrome prevalence and identify prevention strategies.

**Methods:** This was a retrospective cohort study using China Health and Retirement Longitudinal Study (CHARLS) data with a four-year follow-up period (2011--2015), which is a multilevel complex sampling design survey of the Chinese population used to represent the national population. We retrospectively included 4821 participants (27.2% of all participants, mean age = 58 years, 54.1% were female) with sufficient CKM indicator data from CHARLS. We investigated the incidence and progression of CKM syndrome in Chinese adults by building a multivariate logistic regression model to analyze the additional risk factors for CKM progression, focusing on the potential social determinants of health (SDOHs).

**Results:** In the baseline survey, after weighting, the proportions of patients with stages 0-4 CKM syndrome were 10.5%, 17.0%, 46.3%, 12.1%, and 14.1%, respectively. During the 4-year follow-up, 27.20% of patients experienced CKM deteriorated. A higher CRP (OR =1.55, 95% CI: 1.06-2.28, P = 0.02), Chinese visceral obesity index (CVAI) (OR = 1.81, 95% CI: 1.31-2.52, P < 0.001), and conicity index (CI) (OR = 1.34, 95% CI: 1.01-1.79, P = 0.04) were identified as risk factors for CKM deterioration.

**Conclusion:** CKM syndrome incidence is extremely high in middle-aged and elderly people in China, with rapid and severe progression. Additional risk factors and predictive indications related to the staged progression of CKM syndrome should be actively explored to achieve the slowing and reversal of CKM syndrome deterioration.

**Key Points:** This cohort study investigated the prevalence of CKM syndrome and explored the factors related to its progression and recovery.

**Question:** What is the prevalence of CKM syndrome, and what factors affect its progression?

**Findings:** In this cohort study of 4821 participants with 4 years of follow-up, a high incidence of CKM syndrome and a high risk of CKM deterioration were observed in the Chinese middle-aged and elderly population.

**Meaning:** The findings of this study support the recommendations of the AHA that additional risk factors and predictive indicators for the progression of CKM should be actively explored.

## I. Introduction

Cardiovascular-kidney-metabolic syndrome (CKM) is a group of progressive diseases that begin in the young and middle-aged stages of the life cycle. The progression of CKM syndrome is promoted by various risk factors, including inflammation, oxidative stress, and metabolic disorders, leading to a significant increase in cardiovascular disease (CVD) and mortality^1^. The definition and staging of CKM syndrome consider the interaction of multiple chronic diseases, including obesity, diabetes, and kidney disease. The progression of CVDs from dysfunctional obesity (stage 1) and metabolic risk factors (stage 2) to subclinical (stage 3) and even clinical CVDs (stage 4) has important advantages in the prevention of CVDs^2^.

The “CKM health: A Presidential Advisory from the American Heart Association” noted that implementing the CKM conceptual framework for the improvement of global CKM health depends on effective investigations within health centers worldwide while considering the impacts of adverse social determinants of health (SDOHs) in all populations^1^. Healthy lifestyle interventions, especially obesity control, have been found to block or even reverse the progression of CKM syndrome in the early stage^1, 3^. However, in addition to the known metabolic risk factors associated with the CKM concept, few studies have focused on other indicators of CKM progress. To adapt to the current treatment strategy, which combines metabolic risk factor management, renal function protection, and CVD prevention, it is highly important to clarify and optimize the conceptual framework of CKM syndrome and actively identify predictive indicators for the improvement or deterioration of CKM status^4^.

Therefore, the aim of this study was to investigate the incidence of CKM syndrome and analyze disease progression and related risk indicators, with the goal of improving the conceptual framework of CKM and improving CKM health worldwide.

## II. Methods

### 2.1. The population and data source

The cohort data used in this study were from the CHARLS, which focused mainly on people aged 45 years and older. Sampling methods for CHARLS have been described in previous reports^5, 6^. A complex sampling design was adopted to obtain nationally representative data. The baseline data from the CHARLS wave 1 survey included 17705 participants from 450 communities or towns in 150 districts/counties/cities in China. We excluded participants with missing data related to the calculation of the glomerular filtration rate (n = 6091), data on fasting blood glucose (n = 1192), and anthropometric information such as blood pressure (n = 1620). For some participants, data on certain demographic variables (n = 13) and a history of cardiovascular and cerebrovascular diseases (n = 36) were missing. We also excluded patients with missing follow-up data (n = 3932). Ultimately, a total of 4821 participants were included in the study cohort.

The CHARLS approval process for ethical review was conducted by the Beijing University Institutional Review Committee. All study participants provided formal written consent for their participation. The principles outlined in the Strengthening the Reporting of Observational Studies in Epidemiology (STROBE) were followed in this study.

### 2.2. Definition, staging and data analysis of CKM

According to the CKM syndrome concept framework from the American Heart Association, we identified each participant’s CKM syndrome in stages: stage 0 referred to no CKM risk factors, stage 1 included dysfunctional obesity or prediabetes, stage 2 included a high risk of chronic kidney disease (CKD) or metabolic risk factors, stage 3 included a very high risk of CKD or a high predicted 10-year CVD risk, and stage 4 indicated physician-confirmed cardiovascular or cerebrovascular disease^1, 4^. The CKM stage 3 and 4 disease are defined as advanced CKM syndrome^4^. A detailed description of the staging definitions on the basis of the data available in CHARLS is provided in Supplementary Material 1.

### 2.3. CVD and outcome assessment

The main outcome of the study was the incidence of staged deterioration of CKM syndrome, which was defined as patients presenting with a later stage of CKM syndrome at follow-up than at baseline. In contrast, patients with earlier stages of follow-up were defined as having CKM stage recovery, and patients whose stage did not change were referred to as having stable CKM.

We defined patients who entered CKM3 or CKM4 after the baseline survey as the CKM deterioration^4^. Patients with a very high risk of CKD or patients with a 10-year total CVD risk of more than 20% were defined as having subclinical CVD (CKM stage 3). In accordance with the concept of CKM, subjects who self-reported receiving a physician’s diagnosis of heart disease or stroke were defined as having CVD (CKM stage 4).

To improve the cardiovascular health of the population, the achievement of phased recovery in patients with early CKM syndrome was added as a secondary outcome. In patients with stage 1 and 2 CKM syndrome, we investigated factors associated with secondary outcomes (CKM stage 0).

### 2.4. Risk factors and covariates

The following additional risk factors were included: (i) demographics, such as age, sex, and education level; (ii) lifestyle information, such as smoking status, alcohol consumption, sleep duration, and physical activity; (iii) laboratory parameters, such as C-reactive protein (CRP) and uric acid (UA) concentrations; and (iv) metabolism-related indices, such as the Chinese visceral obesity index (CVAI), conicity index (CI), and triglyceride-glucose index (TyG index). Patients aged ≥ 60 years were consideredbe older. The level of education was divided into illiterate, basic education (primary or secondary education), and higher education (high school education or above). Smoking status was defined according to a patient’s self-reported smoking experience. Patients who fit the description of the “I never drink or rarely drink” option were defined as nondrinkers, and all other participants were classified as drinkers (1 or more drinks per month). Sleep duration was self-reported and classified as follows: < 6 h/d was defined as inadequate sleep, and ≥ 9 h/d was defined as excessive sleep^7^. Physical activity status was defined as moderate-intensity physical activity that lasted for 10 minutes or more at least once a week. High CRP and UA levels were defined as higher than the median value among the population. The CVAI is a classification of obesity that reflects the distribution of visceral fat and is calculated via a formula that includes age, sex, waist circumference, BMI, and triglyceride and HDL cholesterol concentrations^8, 9^. The CI reflects the abdominal fat distribution. The TyG index has been widely used to assess obesity insulin resistance^10, 11^. We constructed receiver operating characteristic (ROC) curves to predict the progression of CKM and calculated the areas under the curve (AUCs) and cutoff points.

Additionally, the adverse health social determinants of health (SDOH) risk factors emphasized by the AHA in CKM syndrome, including social participation, depressive episodes, income, health insurance status, and health check-up status, were analyzed in this study^1, 12, 13^. Social participation was confirmed by ten social activities^14, 15^. Depressive episodes were judged by the CES-D10 scale, and a score ≥ 10 was defined as depression^14, 15^. Income was divided into five grades, which were determined by the participants evaluating their family income. Health insurance and health examination status were confirmed via questionnaires.

### 2.5. Statistical methods

In this study, we used CHARLS data from 2011--2015 with a multistage complex sampling design using blood sample weighting and then calculated the incidence of stage 0--4 CKM syndrome.

The proportion and baseline characteristics of participants at each stage of CKM syndrome were calculated as the mean ± standard deviation (SD) of each continuous variable and the number (percentage) of categorical variables. To investigate risk factors for the progression of CKM, multivariate logistic regression was performed. In addition, to determine the risk factors for the progression of CKM syndrome, we investigated the percentage and characteristics of patients with staged deterioration, recovery, and stabilization of CKM syndrome through 2015 follow-up data.

Multivariate logistic regression models were used to assess the associations between risk indicators and new-onset CKM stage 3 or stage 4 (CKM deterioration), and the odds ratios (ORs) and 95% confidence intervals (95% CIs) were calculated. The model included demographics, lifestyle information, laboratory tests, and metabolism-related indices (the CVAI, CI, and TyG) validated by numerous clinical studies. Similarly, we constructed multivariate logistic regression models on the basis of demographics, lifestyle information, and SDOH.

Finally, we established a multivariable logistic regression model for the outcome of achieving stage 0 disease in early CKM syndrome (CKM1 or CKM2) patients, with the model variables including demographic information, lifestyle information, laboratory tests, and CVAI.

All the statistical analyses were performed via R software version 4.3.1 (http://www.r-project.org/), and bilateral P values < 0.05 were considered statistically significant.

## III. The results

### 3.1. Baseline incidence and progression of CKM syndrome

A total of 4821 participants whose mean age was 58 years were included in this study; 54.1% were female (n = 2660). In the 2011 baseline survey, the weighted proportion of patients with CKM syndrome was 10.5% in stage 0, 17.0% in stage 1, 46.3% in stage 2, 12.1% in stage 3, and 14.1% in stage 4. In 2015, the change ratios were 5.8%, 16.1%, 45.4%, 12.7%, and 19.9%, respectively (Figure 1a, Table 1).

**Figure 1.**
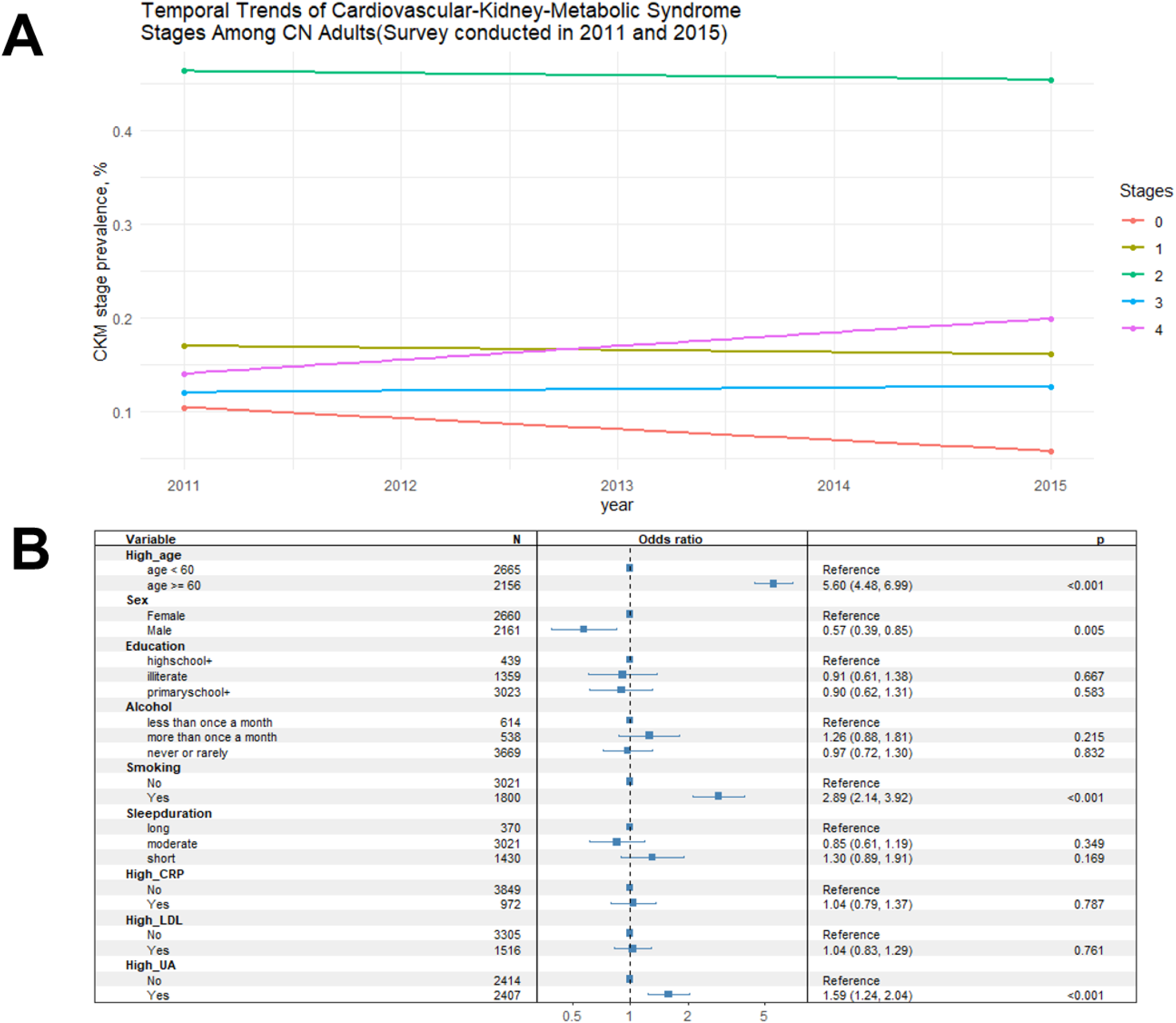
Temporal trends and ORs for CKM syndrome. Panel a-temporal trends in CKM syndrome stages among CN adults in 2011 and 2015. Panel b-Odds ratios for CKM syndrome in multivariate logistic regression. The outcome of the multivariable analysis was the onset of CKM stage 3 or stage 4 (CKM deterioration). with the model variables including age (<60 or ≥60), sex (female or male), education (high school+, illiterate, or primary school+), alcohol (less than once a month, more than once a month, never or rarely), smoking (yes or no), sleep duration (long, moderate or short), high CRP (yes or no), high LDL (yes or no), and high UA (yes or no). High CRP, and UA levels were defined as values higher than the median value among the population. CRP: C­ reactive protein. LDL: low-density lipoprotein. UA: uric acid.

**Table 1.**
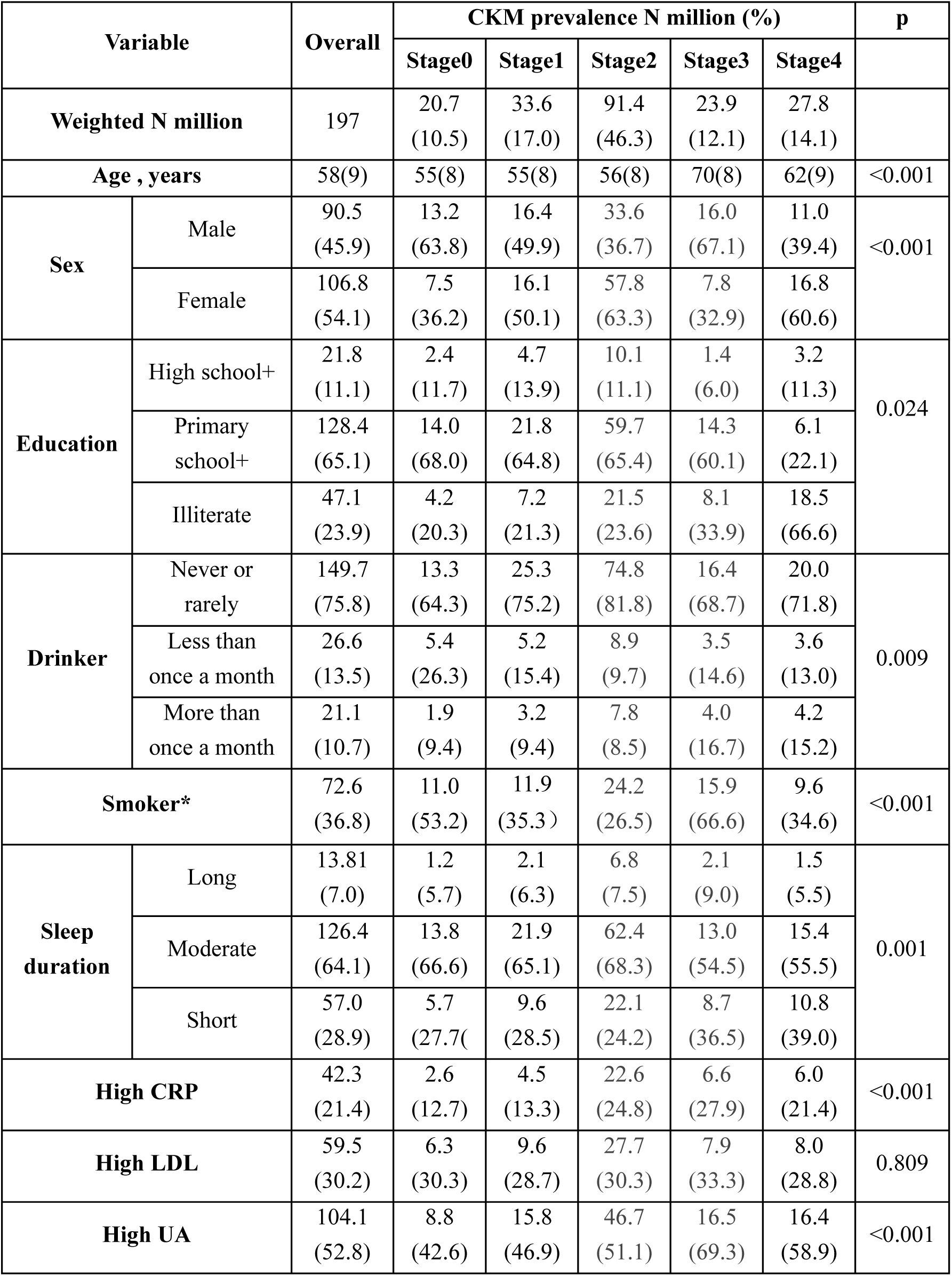
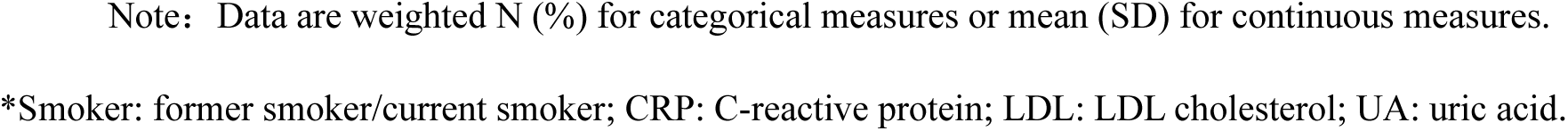
Baseline characteristics of the weighted study population classified by CVH status.

| Variable |  | Overall | CKM prevalence N million (%) |  |  |  |  | p |
| --- | --- | --- | --- | --- | --- | --- | --- | --- |
|  |  |  | Stage0 | Stage1 | Stage2 | Stage3 | Stage4 |  |
| **Weighted N million** |  | 197 | 20.7<br>(10.5) | 33.6<br>(17.0) | 91.4<br>(46.3) | 23.9<br>(12.1) | 27.8<br>(14.1) |  |
| **Age , years** |  | 58(9) | 55(8) | 55(8) | 56(8) | 70(8) | 62(9) | <0.001 |
| **Sex** | Male | 90.5<br>(45.9) | 13.2<br>(63.8) | 16.4<br>(49.9) | 33.6<br>(36.7) | 16.0<br>(67.1) | 11.0<br>(39.4) | <0.001 |
|  | Female | 106.8<br>(54.1) | 7.5<br>(36.2) | 16.1<br>(50.1) | 57.8<br>(63.3) | 7.8<br>(32.9) | 16.8<br>(60.6) |  |
| **Education** | High school+ | 21.8<br>(11.1) | 2.4<br>(11.7) | 4.7<br>(13.9) | 10.1<br>(11.1) | 1.4<br>(6.0) | 3.2<br>(11.3) | 0.024 |
|  | Primary school+ | 128.4<br>(65.1) | 14.0<br>(68.0) | 21.8<br>(64.8) | 59.7<br>(65.4) | 14.3<br>(60.1) | 6.1<br>(22.1) |  |
|  | Illiterate | 47.1<br>(23.9) | 4.2<br>(20.3) | 7.2<br>(21.3) | 21.5<br>(23.6) | 8.1<br>(33.9) | 18.5<br>(66.6) |  |
| **Drinker** | Never or rarely | 149.7<br>(75.8) | 13.3<br>(64.3) | 25.3<br>(75.2) | 74.8<br>(81.8) | 16.4<br>(68.7) | 20.0<br>(71.8) | 0.009 |
|  | Less than once a month | 26.6<br>(13.5) | 5.4<br>(26.3) | 5.2<br>(15.4) | 8.9<br>(9.7) | 3.5<br>(14.6) | 3.6<br>(13.0) |  |
|  | More than once a month | 21.1<br>(10.7) | 1.9<br>(9.4) | 3.2<br>(9.4) | 7.8<br>(8.5) | 4.0<br>(16.7) | 4.2<br>(15.2) |  |
| **Smoker\*** |  | 72.6<br>(36.8) | 11.0<br>(53.2) | 11.9<br>(35.3) | 24.2<br>(26.5) | 15.9<br>(66.6) | 9.6<br>(34.6) | <0.001 |
| **Sleep duration** | Long | 13.81<br>(7.0) | 1.2<br>(5.7) | 2.1<br>(6.3) | 6.8<br>(7.5) | 2.1<br>(9.0) | 1.5<br>(5.5) | 0.001 |
|  | Moderate | 126.4<br>(64.1) | 13.8<br>(66.6) | 21.9<br>(65.1) | 62.4<br>(68.3) | 13.0<br>(54.5) | 15.4<br>(55.5) |  |
|  | Short | 57.0<br>(28.9) | 5.7<br>(27.7) | 9.6<br>(28.5) | 22.1<br>(24.2) | 8.7<br>(36.5) | 10.8<br>(39.0) |  |
| **High CRP** |  | 42.3<br>(21.4) | 2.6<br>(12.7) | 4.5<br>(13.3) | 22.6<br>(24.8) | 6.6<br>(27.9) | 6.0<br>(21.4) | <0.001 |
| **High LDL** |  | 59.5<br>(30.2) | 6.3<br>(30.3) | 9.6<br>(28.7) | 27.7<br>(30.3) | 7.9<br>(33.3) | 8.0<br>(28.8) | 0.809 |
| **High UA** |  | 104.1<br>(52.8) | 8.8<br>(42.6) | 15.8<br>(46.9) | 46.7<br>(51.1) | 16.5<br>(69.3) | 16.4<br>(58.9) | <0.001 |

Note: Data are weighted N (%) for categorical measures or mean (SD) for continuous measures.

Adults ≥ 60 years of age were more likely to develop advanced CKM syndrome (OR = 5.60, 95% CI: 4.48-6.99, P < 0.001). Men were less likely to develop advanced CKM syndrome (OR = 0.57, 95% CI: 0.39-0.85, P = 0.005). Patients with advanced CKM syndrome were more likely to have a history of smoking (OR = 2.89, 95% CI: 2.14-3.92, P < 0.001) and a high Uric acid (OR = 1.59, 95% CI: 1.24-2.04, P < 0.001) (Figure 1b).

Among the 197 million people represented by the sample in this study, the proportions of patients with stage deterioration and recovery of CKM were 27.2% and 12.7%, respectively, while the proportion of patients with stable CKM stage disease was 60.1%. We found that women were more likely to have CKM staged stabilization than those with staged deterioration or recovery. Patients who never smoked were more likely to achieve staged stabilization of CKM (Table 2).

**Table 2.** Baseline characteristics of the weighted study population classified by CVH progression.

| Variable |  | Overall | CKM<br>deterioration | CKM<br>recovery | CKM<br>stabilization | P |
| --- | --- | --- | --- | --- | --- | --- |
| Weighted N million (%) |  | 197.3 | 53.7(27.2) | 25.1(12.7) | 118.5(60.1) |  |
| Age | <60 | 112.0(56.8) | 28.7(53.5) | 14.7(58.6) | 68.6(57.9) | 0.322 |
| | $\geq 60$ | 85.3(43.2) | 25.0(46.5) | 10.4(41.4) | 49.9(42.1) | |
| Sex | Male | 90.5(45.9) | 29.2(54.4) | 14.1(56.1) | 47.2(39.8) | <0.001 |
|  | Female | 106.8(54.1) | 24.5(45.6) | 11.0(43.9) | 71.3(60.2) |  |
| Education | High school<br>+ | 21.8(11.1) | 6.3(11.6) | 2.1(8.4) | 13.4(11.3) | 0.445 |
|  | Primary school + | 128.4(65.1) | 35.0(65.2) | 17.5(69.7) | 75.9(64.0) |  |
|  | Illiterate | 47.1(23.9) | 12.4(23.1) | 5.5(21.9) | 29.2(24.6) |  |
| **Drinker** | Never or rarely | 149.7(75.8) | 39.0(72.6) | 18.0(71.2) | 92.8(78.3) | 0.090 |
|  | Less than once a month | 26.6(13.5) | 9.6(17.7) | 4.1(16.2) | 13.0(11.0) |  |
|  | More than once a month | 21.1(10.7) | 5.2(9.7) | 3.2(12.6) | 12.7(10.7) |  |
| **Smoker\*** |  | 72.6(36.8) | 21.0(39.1) | 12.8(51.0) | 38.7(32.7) | <0.001 |
| **Sleep duration** | Long | 13.8(7.0) | 3.2(5.9) | 1.6(6.5) | 9.0(7.6) | 0.346 |
|  | Moderate | 126.5(64.1) | 35.7(66.4) | 15.4(61.1) | 75.5(63.7) |  |
|  | Short | 57.0(28.9) | 14.8(27.6) | 8.1(32.4) | 34.0(28.7) |  |
| **High CRP** |  | 42.3(21.4) | 11.4(21.3) | 5.5(22.1) | 25.3(21.4) | 0.964 |
| **High LDL** |  | 59.5(30.2) | 17.4(32.4) | 6.7(26.5) | 35.5(30.0) | 0.417 |
| **High UA** |  | 104.1(52.8) | 27.7(51.5) | 13.2(52.7) | 63.2(53.3) | 0.813 |

Note: Data are weighted N million (%) for categorical measures. \*Smoker: former smoker/current smoker; CRP: C-reactive protein; LDL: LDL cholesterol; UA: uric acid.

### 3.2. Additional risk factors and indications for CKM syndrome deterioration

We further explored additional risk factors and indicators for CKM deterioration (new-onset CKM stage 3 or stage 4) in patients with CKM syndrome (Figure 2a). During the 4 years of follow-up, 14.7% of patients experienced CKM deterioration. We found that adults ≥60 years of age, men were more likely to experience CKM deterioration (OR = 5.93, 95% CI: 4.57-7.69, P < 0.001; OR = 2.89, 95% CI: 1.84-4.52, P < 0.001, respectively), whereas smokers were less likely to experience CKM deterioration (OR = 0.64, 95% CI: 0.43-0.98, P = 0.04). We also found that inflammatory factors such as CRP concentrations (OR = 1.55, 95% CI: 1.06-2.28, P = 0.02) were significantly associated with CKM deterioration. Moreover, obesity status and metabolism-related indices were associated with CKM deterioration, with a higher CVAI and CI increasing the deterioration risk significantly (OR = 1.82, 95% CI: 1.31-2.52, P < 0.001; OR = 1.34, 95% CI: 1.01-1.79, P = 0.04). However, we did not observe significant associations between education level, alcohol consumption status, sleep duration, exercise or TyG and CKM deterioration.

**Figure 2.**
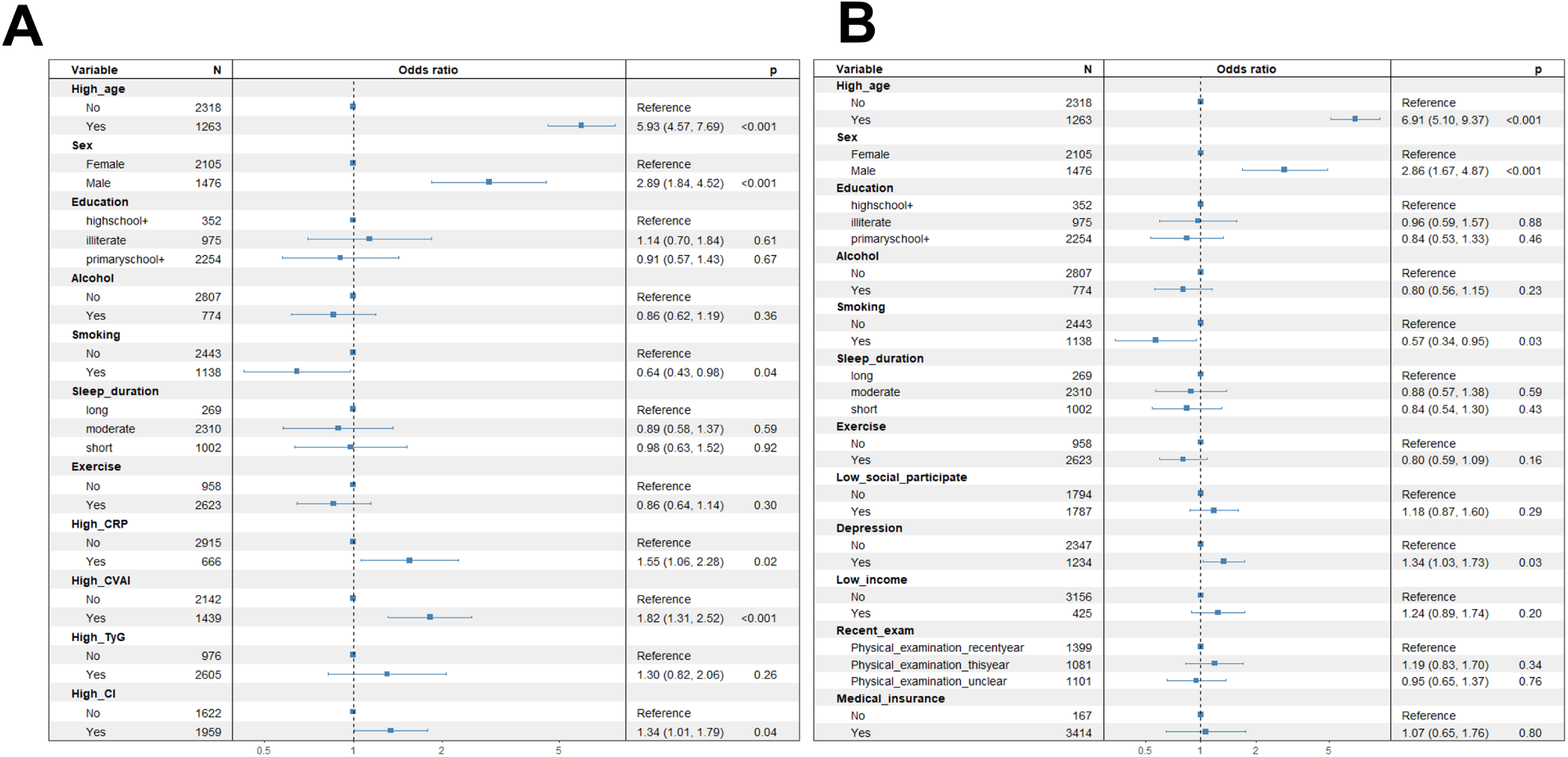
Forest plot of the OR for new CKM deterioration according to multivariate logistic regression. Panel a-The outcome of the multivariable analysis was new onset of CKM stage 3 or stage 4 (CKM deterioration), with the model variables including age (<60 or ≥60), sex (female or male), education (high school+, illiterate, or primary school+), alcohol (less than once a month, more than once a month, never or rarely), smoking (yes or no), sleep duration (long, moderate or short), exercise (yes or no), high CRP (yes or no), high CVAI (yes or no), high Cl (yes or no), and high TyG (yes or no). High CRP status was defined as higher than the median value among the population, whereas high CVAI, Cl, and TyG status was defined as higher than the cutoff point calculated from receiver operating characteristic (ROC) curves with the highest areas under the curve (AUCs). Panel b-The outcome of the multivariable analysis was new onset of CKM stage 3 or stage 4 (CKM deterioration), with the model variables including age (<60 or ≥60), sex (female or male), education (high school+, illiterate, or primary school+), alcohol (less than once a month, more than once a month, never or rarely), smoking (yes or no), sleep duration (long, moderate or short), exercise (yes or no), low social participation (yes or no), depression (yes or no), low income (yes or no), housing status (yes or no), recent physical examination (recent year, this year, or unclear), and medical insurance (yes or no). CRP: C-reactive protein. CVAI: Chinese visceral obesity index. Cl: conicity index. TyG: triglyceride-glucose index.

Concerning the adverse social determinants of health and the stage-specific deterioration of CKM (Figure 2b), after joint adjustment for demographic and lifestyle information, we found that participants with depression had an increased risk of CKM deterioration (OR = 1.34, 95% CI: 1.03-1.73, P = 0.03). However, we did not observe significant associations between other SDOH factors and CKM deterioration in this model.

### 3.3. Protective factors for the staged recovery of CKM syndrome patients

During the 4-year follow-up period, 12.7% of patients with CKM syndrome experienced a staged recovery. The investigation of protective factors that contribute to the recovery of CKM health (Figure 3) revealed that an age above 60 years (OR = 0.49, 95% CI: 0.36-0.66, P < 0.001) and a greater CVAI (OR = 0.43, 95% CI: 0.30-0.62, P < 0.001) would reduced the likelihood of staged recovery. However physical activities increased the likelihood of staged CKM recovery by 80% (OR = 1.91, 95% CI: 1.23-2.65, P = 0.003).

**Figure 3.**
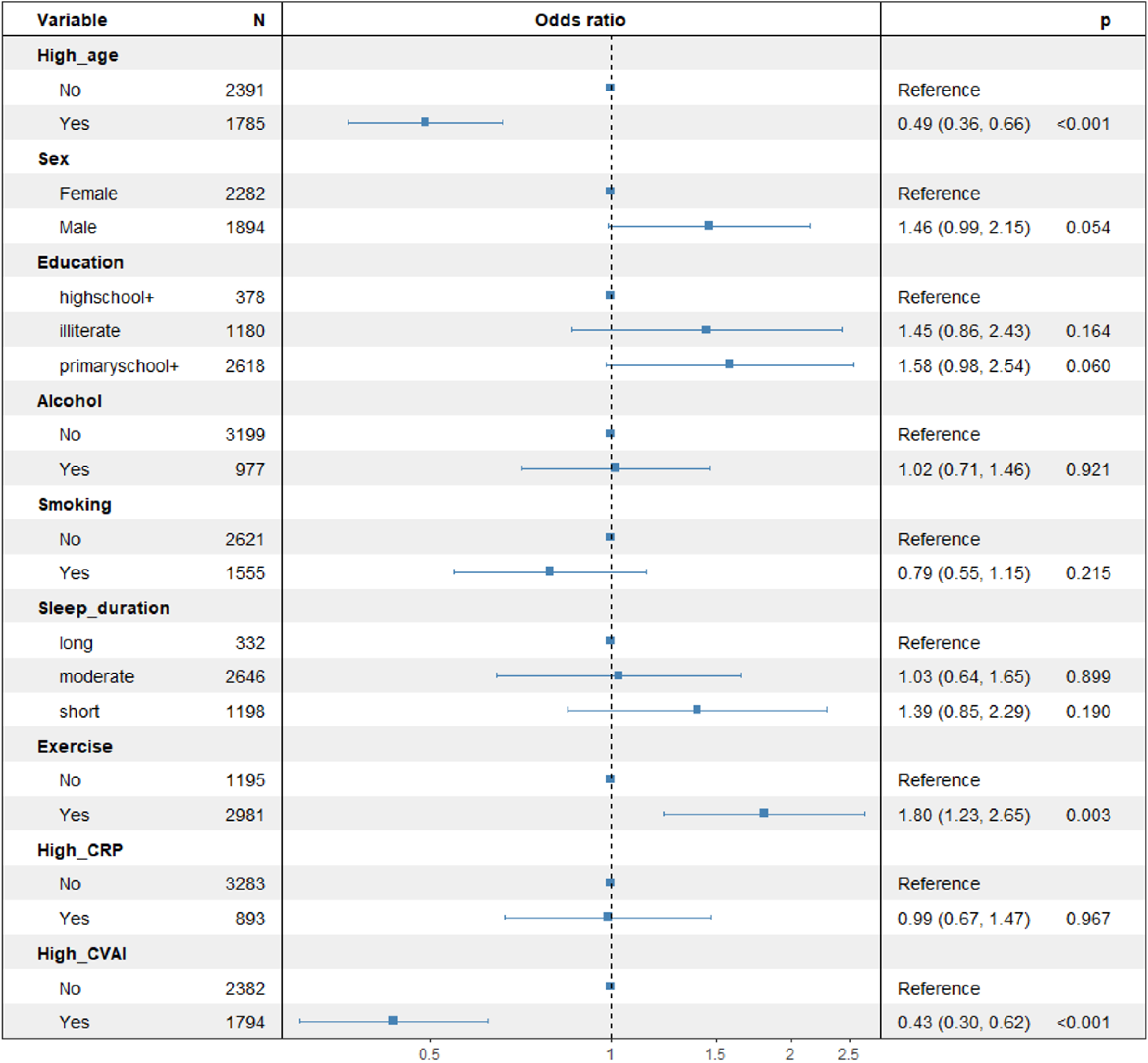
Forest plot of the OR for staged CKM recovery via multivariate logistic regression. The outcome of the multivariable logistic regression model was stage 0 in nonprogressive CKM syndrome patients, with the model variables including age (<60 or ≥60), sex (female or male). education (high school+. illiterate. or primary school+). alcohol (less than once a month, more than once a month, never or rarely), smoking (yes or no), sleep duration (long, moderate or short), exercise (yes or no), high CRP (yes or no). and high CVAI (yes or no). High CRP and CVAI status was defined as higher than the median value among the population. CRP: C-reactive protein. CVAI: Chinese visceral obesity index.

## IV. Discussion

Our results show that the proportion of middle-aged and elderly people in China who meet the diagnostic criteria for CKM syndrome (stage 1 and above) may be close to 90%; in particular, the percentage of patients with advanced CKM syndrome may reach more than 30% (26.2% in 2011, 32.6% in 2015). Second, after our 4-year follow-up survey, the proportion of patients with CKM staged deterioration reached 27.2%. In addition, we found that advanced age, male sex, high CRP level and increased CVAI and CI are additional risk indicators for CKM deterioration, whereas lower age, lower CVAI and exercise are significantly associated with the CKM recovery, which requires more attention in the health management and treatment of CKM to prevent disease deterioration and promote staged recovery.

There is increasing evidence that risk prediction and intervention beginning early in life are important directions for CVD prevention^16, 17^. CKM risk assessment, which is based on the conceptual framework of the CKM, can achieve early prevention of metabolic causes of CVD, and intervention before the occurrence of obvious CVD symptoms can provide maximum benefits to patients’ quality of life^18^. However, the current research on the incidence of multipopulation CKM syndrome as well as the discussion of social determinants of poor health (SODHs) in various regions of the world are still insufficient, which is not conducive to the goal of the CKM conceptual framework regarding the realization of the great value of SODHs for improving global CKM health. In view of this shortcoming, we focused on Chinese people who are already at high risk of CVD and identified the high incidence and rapid deterioration of CKM syndrome^19^. Previous studies of the prevalence of CKM syndrome in the U.S. have reported similar results, with close to 90% of U.S. adults having stage 1 or above CKM syndrome, which supports the findings of this study^4^.

The “CKM Health: The Presidential Advisory from the AHA” recommends that the detection and monitoring of risk factors for CKM should be strengthened to control the worsening of CKM syndrome and prevent irreversible end-stage cardiorenal disease^20–22^. The results of this study indicate that high CRP, and a high CVAI and CI are additional risk indicators for the deterioration of CKM syndrome and have value for predicting the progression of early CKM syndrome, providing evidence for implementing smoking cessation and improving sleep status as early as possible in the life cycle to enhance the primary prevention of CVD^23–25^. We also noted that metabolic and obesity-related indicators have an impact on CKM syndrome via multiple pathways. Abdominal obesity, which is quantified on the basis of the CVAI and Cl, is not only a reliable predictor of metabolic diseases such as metabolic syndrome, nonalcoholic fatty liver, and type 2 diabetes mellitus but also related to independent risk factors such as depression and anxiety status and can serve as a risk indicator for CKM health monitoring^26, 27^. We found that smokers appear to be at lower risk of CKM deterioration, but this may be due to people in good physical condition underestimating the harms of smoking. Although we did not find significant SDOHs other than depression, we recommend that patients during CKM syndrome should be aware of self-care regardless of the availability of healthcare resources and that the provision of healthcare resources for cardiovascular disease needs to be made as equal and accessible as possible.

This study has several strengths. First, this is the first large cohort study to investigate the incidence and progression of CKM syndrome in middle-aged and elderly people in China. Second, on the basis of the follow-up data, this study focused on changes in patients’ CKM stages. For the first time, the staged deterioration of CKM, the phased recovery of CKM, and the stabilization of CKM were taken as outcomes, and the characteristics of populations with different progression outcomes of CKM were analyzed. In addition, we explored multiple additional risk factors outside the existing conceptual framework of CKM in people experiencing CKM progression. This study provides new perspectives and evidence for both global epidemiological studies and conceptual optimization of the CKM.

Several limitations should be considered. First, owing to cohort design reasons or cost considerations, the CHARLS study did not provide urine sample data, so we could only assess CKD status with the eGFR, which may have led to an underestimation of the prevalence of CKD. However, we found that only 2.42% of CKM2 participants and 0.16% of CKM3 participants had staging results involving CKD risk assessment, which ensured the reliability of our results (there was less risk of prevalence being underestimated). Second, some covariates of interest in this study were self-reported, which might lead to potential recall bias. In addition, we analyzed only the incidence of CKM syndrome in 2011 and 2015, and it is unclear whether the resulting trend of progression will continue into the future. Moreover, the CHARLS is a nationally representative cohort study of middle-aged and elderly people in China, so the generalizations of our findings to other regions, ethnicities, and age groups are limited.

## V. Conclusion

A high incidence of CKM syndrome and a high risk of CKM deterioration during the 4-year follow-up were observed in the Chinese middle-aged and elderly population. Therefore, while paying attention to the high-risk metabolic factors in the conceptual framework of CKM, we should actively explore additional risk factors and predictive indicators for the staged deterioration of CKM and formulate relevant public health prevention and treatment strategies to slow and reverse the progression of CKM syndrome.

## Acknowledgement

This research has been conducted using the China Health and Retirement Longitudinal Study (CHARLS) resources. We are grateful to the participants and staff of the CHARLS for their valuable contributions.

## Author contributions

A.C., J.C., Z.Z., and Y.J.designed this study, A.C. performed the data extraction and statistical analysis. Q.H., Y.W., J.C., X.M., L.H., G.W., and Z.W. validated the statistical analysis. A.C. and Q.H. wrote the original draft of the manuscript, which was revised by Y.W., J.C., Z.Z., and Y.J. All authors made great contributions to the manuscript and approved it for submission. Z.Z. and Y.J. are the guarantors of this work and, as much, had full access to all the data in the study and take responsibility for the integrity of the data and accuracy of the data analysis.

## Data availability

Data can be accessed via http://opendata.pku.edu.cn/dataverse/CHARLS.

